# Proinflammatory and cytotoxic CD38^+^HLA-DR^+^ effector memory CD8^+^ T cells are peripherally expanded in human cardiac allograft vasculopathy

**DOI:** 10.1101/2024.12.23.24319590

**Authors:** Yuko Tada, Sujit Silas Armstrong Suthahar, Payel Roy, Vasantika Suryawanshi, Runpei Wu, Erpei Wang, Felix Sebastian Nettersheim, Katarzyna Dobaczewska, Cheryl Kim, Florin Vaida, Gerald P. Morris, Klaus Ley, Paul J. Kim

**Affiliations:** Division of Cardiovascular Medicine, University of California, San Diego, San Diego, CA; La Jolla Institute for Immunology, La Jolla, CA; Immunology Center for Georgia, Augusta University, Augusta, GA; Department of Cardiology, Faculty of Medicine and University Hospital Cologne, University of Cologne, Cologne, Germany; Department of Family Medicine and Public Health, University of California, San Diego, San Diego, CA; Department of Pathology, University of California, San Diego, San Diego, CA

## Abstract

**Background:** T cell mediated immunity is reported to play a pathogenic role in cardiac allograft vasculopathy (CAV) in heart transplant (HTx) patients. However, peripheral blood CD8**^+^** T cells have not been previously characterized in CAV. This study aimed to identify potentially pathogenic circulating CD8^+^ T cell populations in high grade CAV patients using cellular indexing of transcriptomes and epitopes by sequencing (CITE-seq).

**Methods:** Peripheral blood mononuclear cells (PBMC) collected from International Society for Heart and Lung Transplant (ISHLT) grade 2 or 3 CAV (high grade CAV; n=6) and normal HTx (n=12) patients were analyzed using CITE-seq and VDJ-seq. Key findings were validated by flow cytometry in an independent patient cohort of age-matched CAV (n=11) patients, normal HTx (n=12) patients and healthy donor subjects (n=11).

**Results:** Among the seven peripheral CD8**^+^** T cell clusters, high grade CAV patients demonstrated a significantly higher proportion of the CD38**^+^**HLA-DR**^+^** CD8**^+^**effector memory T (Tem) cell cluster compared to normal HTx patients (median 6.2% vs 2.9%, p=0.01). CD38**^+^**HLA-DR**^+^**CD8**^+^** Tem cells showed clonal expansion, activated interferon-γ (IFNG) signaling and enhanced cytotoxicity with granzyme B (GZMB) and perforin (PRF) overexpression. Significantly higher proportion of the proinflammatory and cytotoxic CD38**^+^**HLA-DR**^+^** CD8**^+^**Tem cell cluster in high grade CAV compared to normal HTx patients was validated by flow cytometry. There was significantly increased clonal expansion of peripheral CD8**^+^** T cells in high grade CAV compared to normal HTx patients (median Shannon index = 4.4 vs 6.1, p=0.03). CITE-seq identified LAIR2 as a potential biomarker for identifying high grade CAV patients as increased expression was found in CD38**^+^**HLA-DR**^+^**CD8**^+^** Tem cells. Plasma LAIR2 was significantly elevated in the high grade CAV (n=20) compared to normal HTx patients (n=20; 16.0 pg/mL vs 70.3 pg/mL, p=0.02).

**Conclusions:** We discovered and validated circulating CD38**^+^**HLA-DR**^+^**CD8**^+^** Tem cells to be significantly increased in high grade CAV compared to normal HTx patients. The proinflammatory and cytotoxic phenotype of this CD8**^+^** T cell cluster suggest its potential pathogenic role in human CAV.

**Clinical Perspective:** *What is new?:*

- This is the first study to identify clonal expansion of circulating CD38**^+^**HLA-DR**^+^**effector memory CD8**^+^** T cells in human cardiac allograft vasculopathy.
- CD38**^+^**HLA-DR**^+^** effector memory CD8**^+^** T cells possess both proinflammatory and cytotoxic characteristics, suggesting their potential pathogenic role in human cardiac allograft vasculopathy.
- LAIR2 is a potential signature gene of CD38**^+^**HLA-DR**^+^**effector memory CD8**^+^** T cells.

*What are the clinical implications?:*

- Circulating CD38**^+^**HLA-DR**^+^** effector memory CD8**^+^** T cells and plasma LAIR2 protein are potential early biomarkers of cardiac allograft vasculopathy.
- Evaluation of CD38**^+^**HLA-DR**^+^** effector memory CD8**^+^** T cells in longitudinal studies may reveal how this T cell cluster contributes to the development of human cardiac allograft vasculopathy.
- Inhibiting the expansion of CD38**^+^**HLA-DR**^+^**effector memory CD8**^+^** T cells and/or the LAIR2 pathway may become important therapeutic targets for prevention and treatment of human cardiac allograft vasculopathy.

## Introduction

CAV significantly limits adult HTx longevity to a median survival of 12 years after HTx.^1^ CAV is also the most common reason for re-HTx and responsible for 12% of deaths after 5-y post-HTx. Management of CAV remains challenging even with contemporary immunosuppressive regimens.^2^ Thus, better understanding of CAV immunology may help in earlier detection and development of more effective therapies for CAV.

CAV is widely considered to be mediated by chronic allogeneic immune responses.^3^ However, despite decades of research, the alloimmune mechanisms contributing to human CAV remain largely unresolved. There is evidence from animal models that CD8^+^ T cell mediated immunity plays a central role in the pathogenesis of allograft vasculopathy through the IFNG-axis.^4,5^ In addition, prior investigators have suggested that CD8^+^ T cell responses in chronic rejection are resistant to calcineurin inhibition and capable of mediating CAV by proinflammatory and cytotoxic pathways.^6,7^ However, the role of CD8^+^ T cells in CAV continues to be a subject of controversy, due to the variable mechanisms for allograft vasculopathy in different animal models.^4^

Thus, mechanisms of triggering T cell-mediated immunity in human CAV remain poorly understood. The limited studies of myocardial tissues from CAV patients have previously described prominent infiltration of CD4**^+^** T cells and CD8**^+^** T cells with a Th1 immune response.^3,8^ Peripheral T cells in CAV patients have also not been thoroughly evaluated and limited by flow cytometry panels without strong rationale for the selected antibodies.^9,10^ Thus, we conducted a high-dimensional analysis of PBMCs from high grade CAV and normal HTx patients using CITE-seq and T cell receptor sequencing (TCR-seq) with subsequent validation of our findings using flow cytometry in an independent HTx patient cohort. We sought to test the hypothesis that circulating IFNG expressing CD8**^+^** T cells differentiate and expand in HTx patients to contribute to developing high grade CAV.

## Methods

### Data Availability

The data will be made available to other researchers for purposes of reproducing the results upon request. A detailed Materials and Methods section is available in the Supplemental Material.

### Independent data access and analysis

The corresponding author (PJK) has full access to all the data and takes responsibility for the integrity and data analysis.

### Cohort Description

This study was approved by the University of California, San Diego (UCSD) Health Office of IRB administration (No. 160808), and all participants provided written informed consent. Eligible patients were HTx recipients who were 18 y of age or older. Exclusion criteria were HTx from hepatitis C nucleic acid test positive donors and patients with a previous HTx. CAV was diagnosed by the angiographic findings and graded according to the ISHLT criteria.^11^ HTx patients diagnosed with ISHLT grade 2 or 3 CAV were recruited in the high grade CAV group. HTx patients with normal graft function (normal HTx) were defined by no presence of CAV by coronary angiography, dobutamine stress echo, or stress perfusion MRI in addition to LV ejection fraction (LVEF) higher than 50% and no symptoms of heart failure. High grade CAV and normal HTx patients were frequency matched for age, gender, race, time after HTx, and multi-organ transplant status for CITE-seq and flow cytometry. Young healthy donors (<50 years old and without HTx) were recruited through the Normal Blood Donation Program at the La Jolla Institute for Immunology. Older healthy donors (≥50 years old and without HTx) were recruited from those who had a 0 coronary calcium score by screening at UCSD Health. Young and older healthy donors thus contributed to the healthy control (HC) group and were also screened for other comorbidities, including uncontrolled hypertension, dyslipidemia, diabetes mellitus, and chronic viral infections such as hepatitis B, hepatitis C, and HIV.

### Statistical Analysis

Data were expressed as median and IQR. Statistical analysis was performed using R (R Core Team, 2024) and GraphPad Prism (v10). Statistical comparison of numerical variables between the two groups were performed with the Wilcoxon rank-sum test. Multiple group comparisons were performed with Kruskal-Wallis test followed by pairwise group comparisons using the Dunn multiple-comparisons test. Statistical comparisons of categorical variables were performed using Fisher’s exact test. For paired comparisons of median fluorescence intensity (MFI) data from flow cytometry between the cell types within each patient, paired non-parametric tests (Friedman test) were performed and p-values were adjusted by the Bonferroni-Holm procedure. Adjusted or unadjusted p values were designated as p_adj, or p, respectively. P or p_adj <0.05 were considered significant.

## Results

### Major peripheral immune cell types using CITE-seq

The design of this study is shown in **Figure 1A**. We analyzed PBMC samples collected from normal HTx (n=12) and high grade CAV (n=6) patients using CITE-seq and TCR-seq. Demographics and clinical parameters of the patients are summarized in **Table S5**. Compared to the normal HTx group, patients in the high grade CAV group showed significantly lower left ventricular ejection fraction. Prevalence of hypertension and diabetes was equivalent in both groups. Two high grade CAV patients died, and 4 high grade CAV patients had a re-HTx during the follow-up period (2021-2024) while no normal HTx patients experienced death or re-HTx.

**Figure 1.**
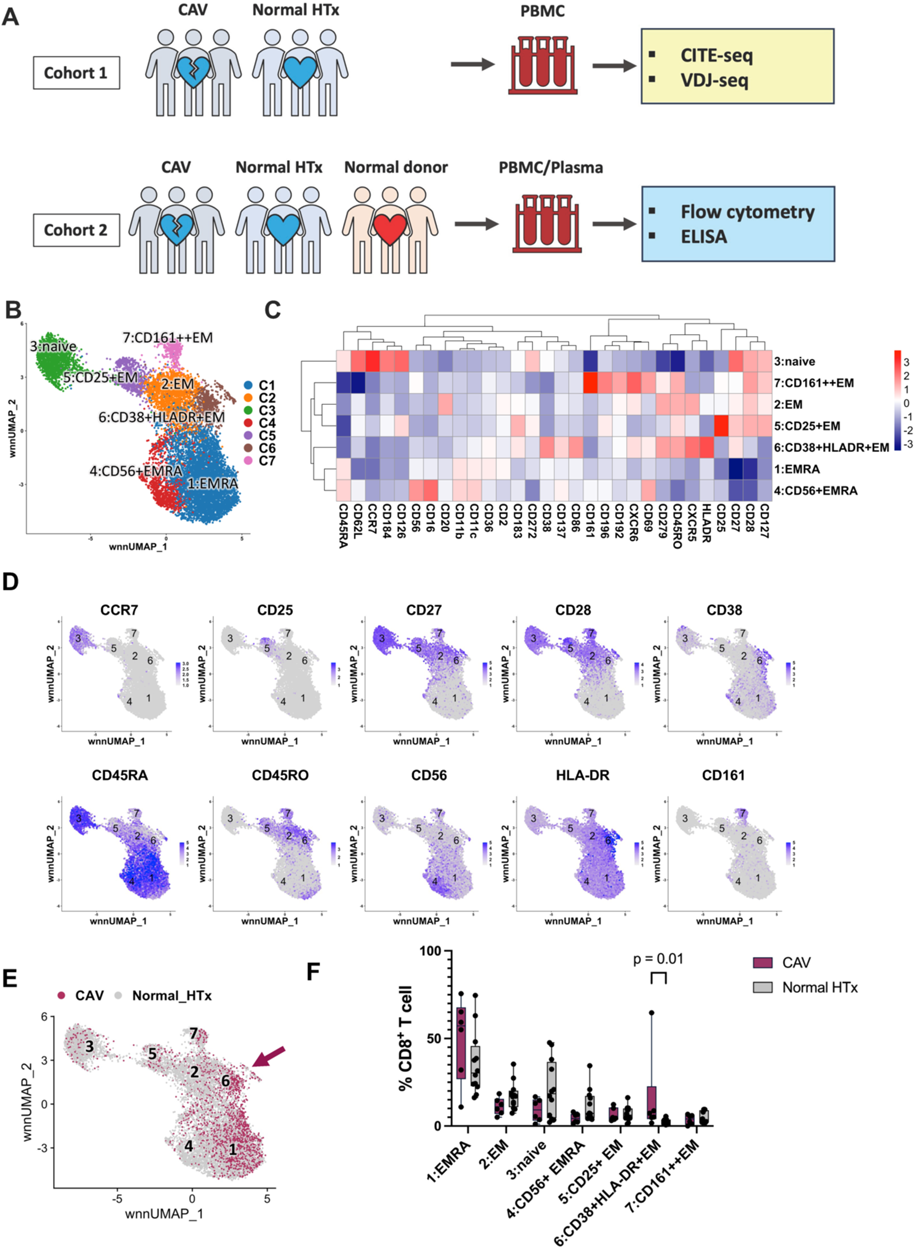
Analysis of peripheral CD8^+^ T cell clusters by CITE-seq reveals expansion of CD38^+^HLA-DR^+^ CD8^+^ Tem cells in high grade CAV patients. **A.** Schematic representation of the experimental design. Cohort 1 included the high grade CAV and normal HTx patient groups. Cohort 2 included high grade CAV, normal HTx, and normal donor groups. Peripheral blood mononuclear cells (PBMC) obtained from cohort 1 were used for single cell CITE-seq and VDJ-seq. PBMC and plasma obtained from cohort 2 were used for flow cytometry and ELISA. **B.** Circulating CD8^+^ T cell clusters were identified across all samples (n=18) and displayed on the UMAP. **C.** Heatmap shows expression patterns of ADT among the CD8^+^ T cell clusters. Color scale represents the scaled values of centered log ratio normalized expression. **D.** Expression of the 10 key surface markers that differentiate CD8^+^ T cell clusters are shown on the UMAPs. **E.** Cells from the high grade CAV vs normal HTx groups are annotated on the UMAP. Cells from the high grade CAV group showed a concentrated distribution in CD38^+^HLA-DR^+^ CD8^+^ Tem cell cluster (C6; arrow). **F.** Proportions of each cluster (% total CD8^+^ T cells) were compared between the high grade CAV and normal HTx groups demonstrating a significantly increased proportion of C6 in high grade CAV (Wilcoxon rank-sum test).

Dimensionality reduction and clustering of the major immune cell types was performed using a final set of 18 ADT markers (**Figures S1** and **S2**). There were 10 major immune cell type clusters identified: naive CD4^+^ T cells, memory CD4^+^ T cells, naive CD8^+^ T cells, memory CD8^+^ T cells, B cells, natural killer (NK) cells, natural killer T (NKT) cells, classical monocytes, nonclassical monocytes, and dendritic cells (DC; **Excel file S1**, **S2**). Surface marker expressions of the major cell types are shown in **Figure S2**.

### Naive peripheral CD4^+^ T cells in high grade CAV compared to normal HTx patients

We analyzed peripheral CD4^+^ T cells to compare our findings with prior literature.^9,10^ Naive (4,103 cells) and memory (8,467 cells) CD4^+^ T cells, and CD4^+^ NKT cells (139 cells) were combined and clustered by weighted nearest neighbor (WNN) analysis.^12^ Nine CD4^+^ T cell clusters were identified based on their surface protein and differential gene expressions (**Figure S3A-C**, **Excel file S3**; differential gene expression). The high grade CAV group showed a trend towards decreased naive population compared to the normal HTx group, likely a result of the differentiation of the peripheral CD4^+^ T cells (**Figure S3D**). However, we did not detect any other trends in cell proportions between the patient groups.

### Circulating CD38^+^HLA-DR^+^ CD8^+^ Tem cells were significantly higher in high grade CAV compared to normal HTx patients

For the analysis of CD8**^+^** T cells, naive (1,696 cells) and memory (10,679 cells) CD8**^+^** T cell clusters were combined. After removal of double positive (CD4**^+^**CD8**^+^**) and double negative (CD4^-^CD8^-^) clusters (**Figure S4A**), the remaining CD8**^+^** T cells (12,375 cells) were analyzed using WNN to identify seven CD8**^+^** T cell clusters (**Figures 1B-D**).

Clusters were called based on the ADT surface marker expressions. Cluster 1 was called effector memory re-expressing CD45RA (Temra; CCR7-CD27-CD45RA+). Cluster 4 also expressed CD56 and was called CD56**^+^**Temra. Cluster 3 was called naïve (CCR7+CD27+CD45RA+). Cluster 2 was called effector memory (Tem; CCR7- CD27+CD45RO+). Cluster 5 also expressed CD25 and was called CD25**^+^**Tem. Cluster 6 was called CD38**^+^**HLA-DR**^+^** Tem because of its expression of the activation markers HLA-DR and CD38. Cluster 6 downregulated CD28 expression compared to Clusters 2 and 5, compatible with its activated status.^13^ Cluster 7 expressed a high level of CD161 as well as CCR2, CCR6 and CXCR6, and was called CD161**^++^** Tem.

Cells from the high grade CAV group were concentrated in Temra (C1) and CD38**^+^**HLA-DR**^+^** Tem (C6) clusters (**Figure 1E**). The proportion of the CD38**^+^**HLA-DR**^+^**Tem cluster (%CD8**^+^** T cell) was significantly increased in the high grade CAV patients (median 6.2%; IQR, 4.8%-8.3%, p=0.01) compared to normal HTx (median 2.9%; IQR, 1.2%-3.6%; **Figure 1F**). Numbers of the CD8**^+^** T cells in each cluster per individual patient are shown in **Figure S4B** and **Excel file S1**.

### CD38^+^HLA-DR^+^ CD8^+^ Tem cells showed increased expression of proinflammatory and cytotoxicity markers compared to other peripheral CD8^+^ T cell clusters

Predicted functions of the CD38**^+^**HLA-DR**^+^**Tem and other CD8**^+^** T clusters were evaluated by differential gene expression (**Figure 2A, S4C, Excel file S3**) and gene set enrichment analysis (GSEA). C3 was compatible with the naive phenotype overexpressing genes involved in cell homeostasis, proliferation, and lymph node homing (CCR7, PASK, LEF1, SELL, PIK3IP1, CD27, TXK, NT5E, and MYC; **Figure 2A**). CD161**^++^** CD8**^+^** Tem cells (C7) overexpressed genes compatible with previously reported CD161 bright Tc17-like CD8**^+^**T cells or membrane associated invariant T cells (KLRB1, RORC, IL18RAP, IL18R1, IKZF2, ZBTB16, and CCR6).^14^ CD25**^+^** Tem (C5) overexpressed activation markers (IL2RA, CD69, CXCR3, and CCR6). The CD8**^+^** Tem cluster (C2) overexpressed genes including GZMK, EOMES, RGS1, and DUSP2. Temra clusters (C1 and C4) overexpressed cytotoxic enzymes (PRF1, GNLY, GZMB). Temra (C1) overexpressed genes involved in the cellular immunity (IFNG, TBX21, NKG7). CD56**^+^** Temra overexpressed FCGR3A and NK-like genes (TYROBP, KLRC3, KLRC1, KLRF1, KIR2DL1).

**Figure 2.**
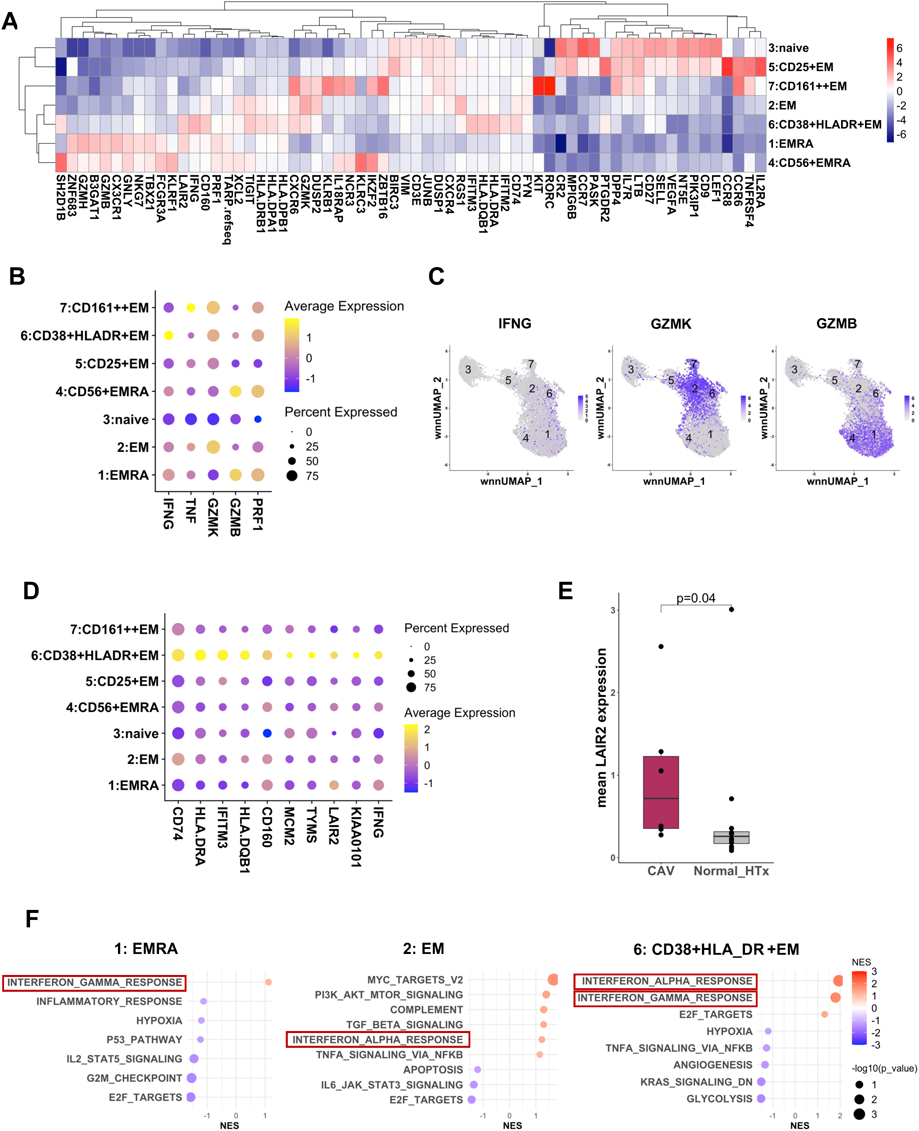
Transcriptomic analysis shows upregulation of inflammatory and cytotoxic genes in circulating CD38^+^HLA-DR^+^ CD8^+^ Tem cells. **A.** The heatmap compares most significantly overexpressed genes among the CD8^+^ T cell clusters. Color represents log2 scaled values. **B.** The dot plot compares expression of inflammatory and cytotoxic genes (IFNG, TNF, GZMK, GZMB, PRF1) across clusters. **C.** Differential expressions of IFNG, GZMK, and GZMB are shown in the featured UMAPs. CD38^+^HLA-DR^+^ CD8^+^ Tem cell cluster (C6) shows the upregulated IFNG expression and intermediate expression of GZMK and GZMB compared to Tem and Temra. **D.** The dot plot shows the expression of potential signature genes of C6 determined as the top 10 most highly expressed genes. **E.** The mean LAIR2 expression in CD8^+^ T cells was significantly increased in the high grade CAV group compared to normal HTx. **F.** Gene set enrichment analysis compared the enriched pathways in the Temra (C1), Tem (C2), and C6, and showed the activation of IFNA and IFNG responses in C6. The dot scales indicate the normalized enrichment score (NES) and p-values.

The CD38**^+^**HLA-DR**^+^** CD8**^+^**Tem cluster overexpressed MHC class II genes (HLA.DRB1, HLA.DRA, HLA.DPA1, HLA.DQB1, HLA.DMA, CD74) and type I IFN- responsive genes (IFITM3, IFITM2; **Figure 2A, S4C**). This cluster also expressed immune checkpoint genes (CD160, TIGIT, and CTLA4). In addition to expressing IFNG, CD38**^+^**HLA-DR**^+^** Tem cluster also expressed higher levels of the cytotoxic enzymes GZMB and PRF1, with a lower expression of GZMK compared to the Tem cluster (C2; **Figures 2B** and **2C**). These results suggest intermediate differentiation of CD38**^+^**HLA-DR**^+^**Tem in between the Tem and Temra clusters.^15^ Among the top 10 most highly overexpressed genes (**Figure 2D**), we discovered LAIR2 as a potential candidate biomarker for high grade CAV. LAIR2 expression was increased in CD8**^+^** T cells from high grade CAV compared to normal HTx patients (**Figure 2E**, **Figure S4D, Excel file S5**).

Activated pathways determined by GSEA were compared between Temra, Tem, and CD38**^+^**HLA-DR**^+^** Tem clusters (**Figure 2F**). CD38**^+^**HLA-DR**^+^** Tem showed the most activated interferon-α (IFNA) and IFNG responses, consistent with the activation of IFNG-axis in CAV.

### CD38 and HLA-DR expression in circulating CD8^+^ T cells

We inferred the differentiation trajectories across CD8**^+^** T cell clusters by analyzing the diffusion map (**Figure 3A**). Diffusion pseudo-time was determined by ranking cells ordered along the inferred diffusion trajectories (**Figure S5A**). Cells in the CD38^+^HLA- DR^+^ Tem cluster (C6) were shown to be between the Tem (C2) and Temra (C1) clusters on the diffusion map (**Figure 3A**). CD8^+^ Temra cells had the highest diffusion pseudo-time (**Figure 3B, C**). Intermediate diffusion pseudo-time of C6 suggested a differentiation trajectory from Tem to CD38^+^HLA-DR^+^ Tem and then to Temra cells. Clonal expansion initially correlated linearly with higher diffusion pseudo-times and peaked at the midpoint of the plotted diffusion pseudo-times (**Figure 3D**). This may be attributable to the contraction of clonal expansion due to the decreased proliferative potential and longevity of terminally differentiated CD8^+^ T cells.^16^

**Figure 3.**
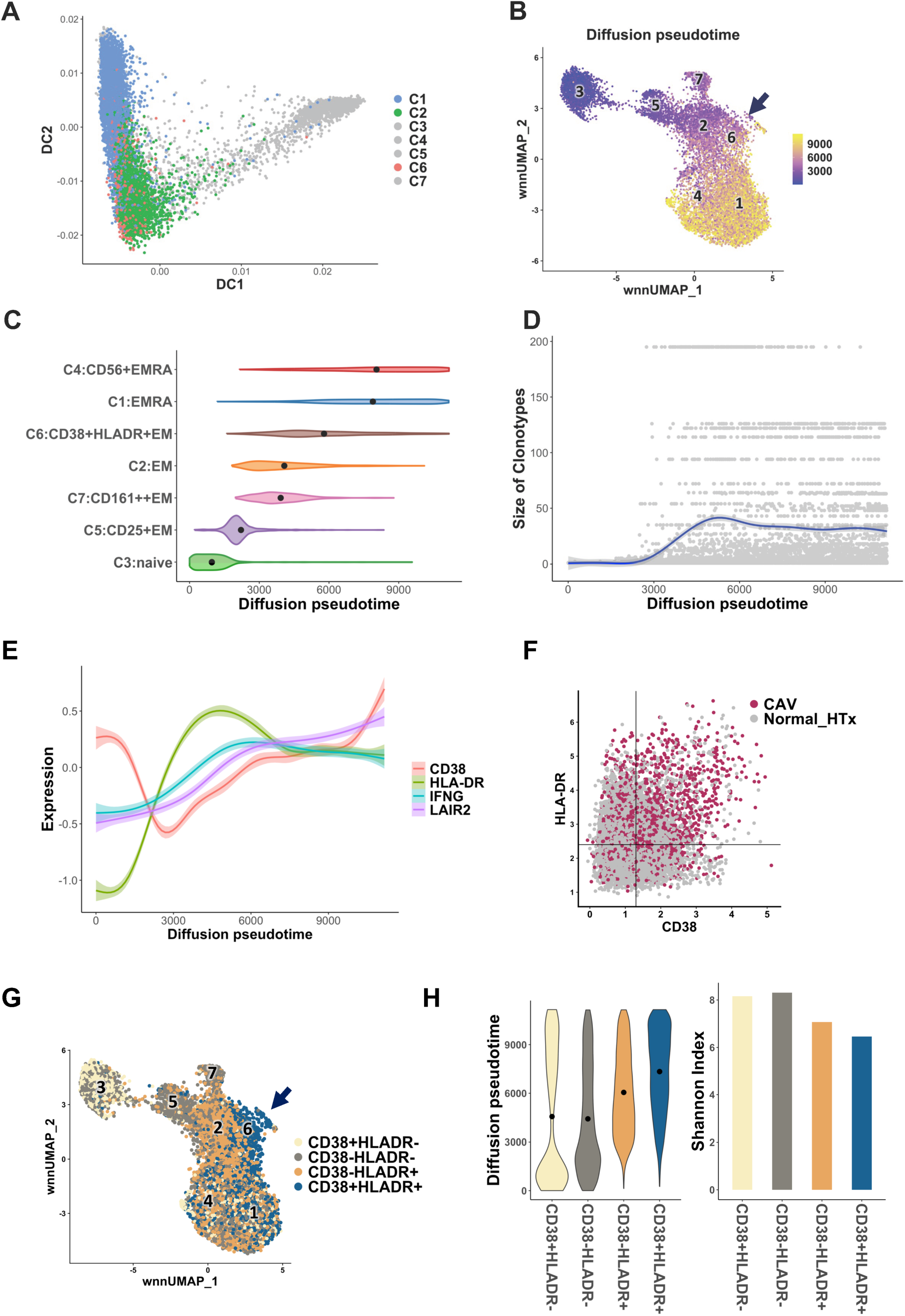
CD38 and HLA-DR expression in circulating CD8^+^ T cells. **A.** The diffusion map was analyzed and cells were plotted across diffusion components 1 (DC1) and 2 (DC2). Cells from Temra (C1), Tem (C2), and CD38^+^HLA- DR^+^ Tem (C6) clusters are colored to show the proximity of C6 with C1 and C2. **B.** The diffusion pseudo-time determined as the ranks of cells ordered along the diffusion trajectories is annotated on the UMAP. C6 pointed by an arrow had diffusion time higher than C2 but lower than C1. **C.** The violin plot compares diffusion pseudo-time among the CD8^+^ T cell clusters. Mean values are represented by the filled circles. **D.** All CD8^+^ T cell clonotypes were plotted across diffusion pseudo-time and the size of clonotypes. The smoothed trend line, representing mean size of clonotypes, peaked at the midpoint of the plotted diffusion pseudo-times. **E.** Trend lines of expression of CD38 protein, HLA-DR protein, IFNG gene, and LAIR2 gene were plotted showing the patterns of their expression related to CD8^+^ T cell differentiation. The y-axis shows the scaled expression. **F.** The scatter plot with a quadrate gate shows concentrated distribution of CD8^+^ T cells from high grade CAV patients in the CD38 and HLA-DR double positive segment. **G.** Different CD38 and HLA-DR expression groups are annotated on the UMAP, which show a recognizable pattern related to CD8^+^ T cell differentiation. C6 consisted of CD38(+)HLA-DR(+) cells (black arrow). **H.** The violin plot (left) and the bar plot (right) compare diffusion pseudo-times and T cell receptor (TCR) Shannon diversity index among the four different CD38 and HLA-DR expression groups, respectively. CD38(+)HLA-DR(+) CD8^+^ T cells show the highest pseudo-time and lowest TCR diversity, consistent with its differentiated and clonally expanded status.

CD38 and HLA-DR surface protein expression plotted against diffusion pseudo-time showed distinct patterns during CD8^+^ T cell differentiation (**Figure 3E**). Upregulation of HLA-DR preceded that of CD38. HLA-DR was downregulated in CD8^+^ T cells with the highest pseudo-time, but CD38 was further upregulated in these cells. Upregulation of IFNG and LAIR2, the potential signature genes of the CD38^+^HLA-DR^+^ CD8^+^ Tem cells, were associated with the expression of CD38 and HLA-DR (**Figure 3E**). However, the expression of IFNG was decreased in the CD8^+^ T cells with the highest pseudo-time, and LAIR2 was further increased suggesting possible association of high LAIR2 expression with CD8^+^ T cell terminal differentiation.

We also compared CD38 and HLA-DR surface protein expression patterns among the clusters by thresholding their expression, which provided us information to guide assessment of T cells by flow cytometry. Thresholds were determined based on the expression by major cell types known to express CD38 (NK cells) and HLA-DR (monocytes, DC, B cells; **Figure S5B**). On the scatter plot, cells from the high grade CAV group were predominantly located in the CD38(+)HLA-DR(+) segment (**Figure 3F**). The UMAP showed recognizable CD38 and HLA-DR expression patterns associated with CD8^+^ T cell differentiation (**Figure 3G**). CD38(+)HLA-DR(+) cells showed the highest pseudo-time and lowest TCR diversity, consistent with their highly differentiated and clonally expanded status (**Figure 3H**). However, the bimodal peak of diffusion pseudo-time was noted in the CD38(+)HLA-DR(-) CD8^+^ T cells (**Figure 3H**). Our data suggest that this finding is due to both naïve and most differentiated CD8^+^ T cells being CD38 positive but HLA-DR negative.

### Circulating CD38^+^HLA-DR^+^ CD8^+^ Tem cells demonstrated clonal expansion and reduced TCR diversity

Clonality of CD8**^+^** T cells was compared between the high grade CAV and normal HTx groups and also among different CD8**^+^** T cell clusters. Diversity of the CDR3αβ clonotypes was assessed by the Shannon diversity index. Different sample sizes were adjusted by calculating the asymptotic diversity using the bootstrapped rarefaction/exploration curves (**Figure S6A**). High grade CAV patients demonstrated significantly decreased CD8**^+^**T cell receptor diversity (median 4.4; IQR, 3.8-4.9; p=0.03) compared to normal HTx (median 6.1; IQR, 5.7-6.6), consistent with increased clonal expansion of peripheral CD8**^+^** T cells (**Figure 4A)**.

**Figure 4.**
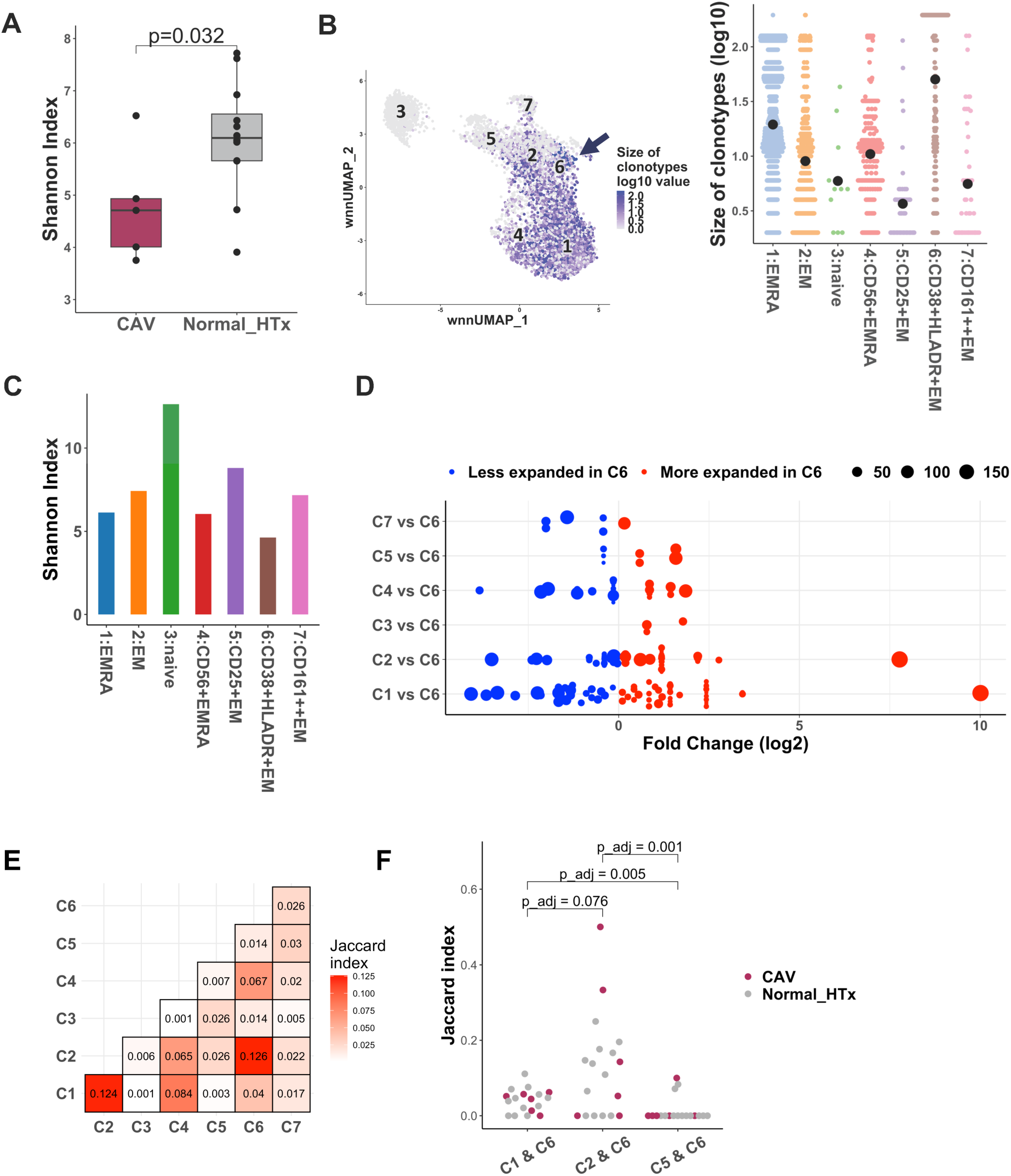
Circulating CD38^+^HLA-DR^+^ CD8^+^ Tem cells demonstrate clonal expansion and increased differentiation. **A.** The Shannon Index showed significantly decreased CD8^+^ T cell T cell receptor (TCR) repertoire diversity in high grade CAV patients compared to normal HTx (Wilcoxon rank-sum test). **B.** The size of clonotypes (log10 scale) are annotated on the UMAP (left) showing the distribution of large clonotypes in the CD38^+^HLA-DR^+^ CD8^+^ Tem cell cluster (C6). The size of expanded clonotypes (log10 scale) were compared among the CD8^+^ T cell clusters (right). The filled black circles represent the mean values. **C.** The Shannon diversity index calculated for each cluster showed the lowest TCR repertoire diversity in C6. **D.** Clonotypes expanded in C6 and shared with other CD8^+^ T cell clusters were filtered and similarities of the clonotypes were compared among the clusters. Shared clonotypes were plotted across relative proportion of cells in C6 vs non-C6 cluster (log2 fold change). Size of each dot represents the size of each clonotype. Proportions of the cells were normalized by the size of the clusters. The shared clonotypes are predominantly found in the Temra (C1) and Tem (C2) clusters. **E.** The heat map shows the Jaccard index comparing the similarities of clonotypes between the clusters. The values are the means of the Jaccard similarity index determined for each sample (n=18). **F.** Clonotype similarities between C1 & C6 and C2 & C6 were significantly higher compared to the similarity between CD25^+^ Tem (C5) & C6 clusters (Wilcoxon rank-sum test, p values adjusted by the Bonferroni-Holm procedure). The filled circles represent the Jaccard similarity index from each patient sample (n=18).

Size of the clonotypes, an indicator of the extent of clonal expansion, was compared among the CD8**^+^** T cell clusters (**Figure 4B**). The CD38^+^HLA-DR^+^ CD8**^+^** Tem cell cluster (C6) showed the largest average size of expanded clonotypes, which was followed by the CD8**^+^**Temra clusters. Consistently, C6 showed the lowest TCR diversity (**Figure 4C**). We evaluated the similarity of C6 with the other CD8**^+^** T cell clusters based on the overlapping TCR repertoires. Clonotypes of C6 were predominantly shared by the Temra (C1) and Tem (C2) clusters (**Figure 4D**). Clonotype similarities were also evaluated by Jaccard index. Mean Jaccard index showed the clonotypes of C6 had the highest similarity with C2, which was followed by C1 and C4 (**Figure 4E**). Similarities between C6 vs C2 and C6 vs C1 were significantly higher compared to C6 vs CD25**^+^** Tem (C5; **Figure 4F**). Clonotype similarities between C6 and all other CD8**^+^** T cell clusters were shown in Figure **S6B**. These results suggest expansion of C1, C2, and C6 by chronic stimulation of the same antigen(s) and was consistent with the posited differentiation trajectory of C6 from C2 into C1.

### Flow cytometry validated significantly increased circulating CD38^+^HLA-DR^+^ CD8^+^ Tem cells in high grade CAV patients

Flow cytometry was performed on PBMCs collected from an independent cohort of high grade CAV patients, normal HTx patients, and HC subjects (**Table S7**). The gating strategy for the major cell types and obtained cell proportions are shown in **Figure S7** and **Table S8**. The CD4/CD8 T cell ratio was significantly decreased in the normal HTx and high grade CAV patients compared to HC (**Figure S7C**).

We gated CD8^+^ T cells using cell surface markers CD161, CD27, CCR7, CD38, HLA-DR and CD56 to reproduce the CITE-seq clusters (**Figure 5A)**. Correspondence of CD8^+^ T cell clusters determined by gating of cell surface markers compared to WNN is summarized in **Tables S9** and **10**.

**Figure 5.**
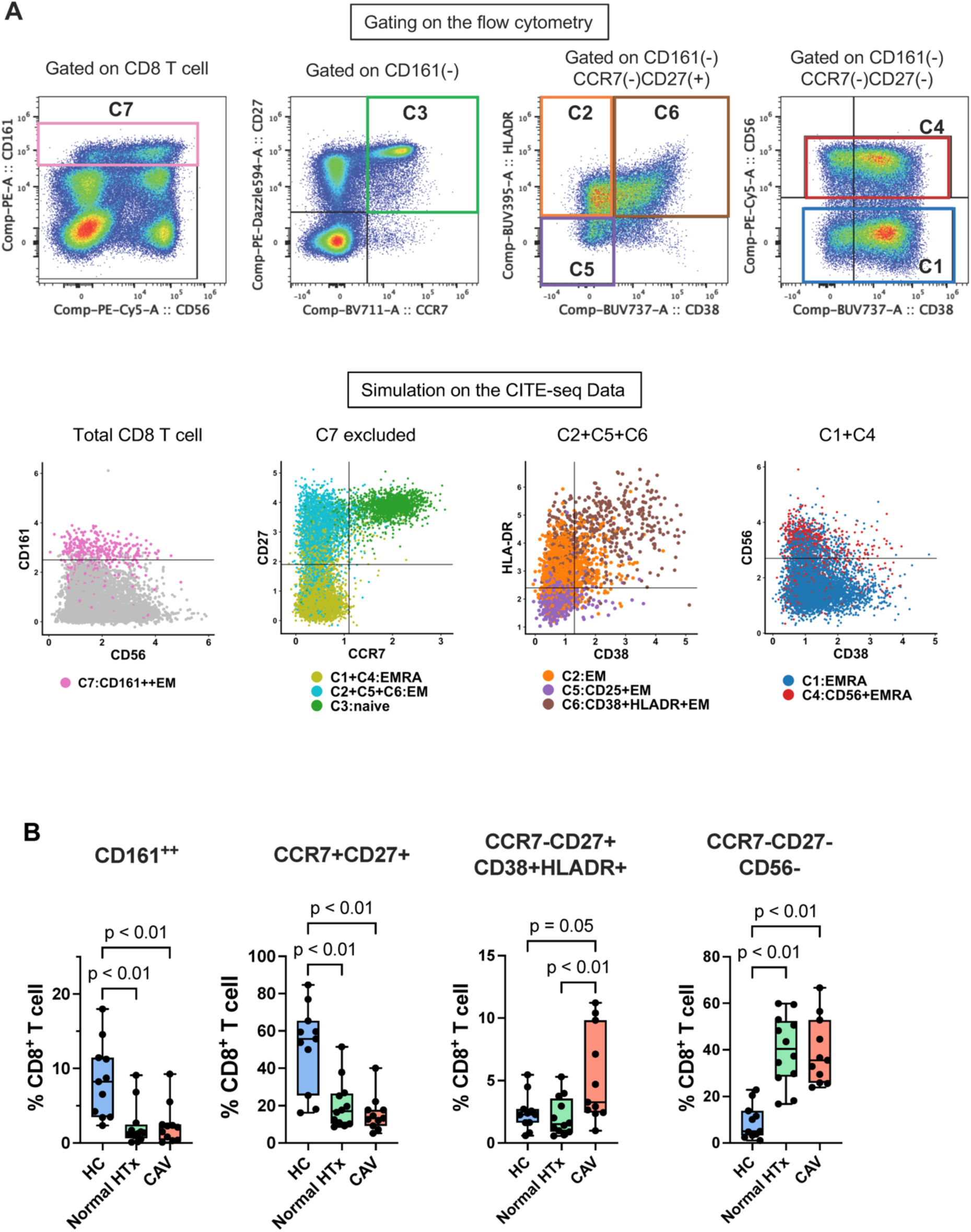
Flow cytometry demonstrates an increased proportion of circulating CD38^+^HLA-DR^+^ CD8^+^ Tem cells in high grade CAV patients. **A.** Gating of CD8^+^ T cells in flow cytometry analysis was determined by simulating on the CITE-seq data to reproduce their clusters. Plots in the upper row show representative gates on the flow cytometry data. CD161^++^ CD8^+^ T (equivalent to C7) was gated first (left). CCR7 vs CD27 gating separated the non-CD161^++^ CD8^+^ T cells into naive, Tem, and Temra clusters (second from left). CCR7-CD27+ Tem clusters were separated into C2, C5, and C6 by CD38 and HLA-DR expression (second from right). CCR7-CD27- Temra cells were gated by CD56 to separate C1 and C4 (right). Plots in the lower row show the simulated gates on the CD8^+^ T cells from CITE-seq. **B.** Proportions of the gated CD8^+^ T cell sub-populations (%CD8^+^ T cells) are compared among high grade CAV, normal HTx, and healthy control (HC) groups. CD38^+^HLA- DR^+^ CD8^+^ Tem cells (C6) were significantly increased in the high grade CAV group compared to normal HTx. Non-adjusted p values <0.05 (Dunn test) are shown.

The proportion of the CD38^+^HLA-DR^+^ CD8^+^ Tem cells was significantly increased in high grade CAV (median 3.3%; IQR, 2.4-9.8%; p=0.009) compared to normal HTx patients (median 1.5%; IQR, 0.9-3.6%), which confirmed our initial CITE-seq findings (**Figure 5B**).

Both high grade CAV and normal HTx patients demonstrated significantly decreased CD161^++^CD8^+^ and naïve CD8^+^ T cell populations (%CD8^+^ T), and significantly increased Temra populations compared to HC. These results suggest increased CD8^+^ T cell activation and potentially immune senescence in HTx patients.^17,18^

### Circulating CD38^+^HLA-DR^+^ CD8^+^ Tem cells expressed both proinflammatory and cytotoxic markers

Expression of IFNG, TNF, GZMK, GZMB, and PRF was compared by the median fluorescence intensity (MFI) obtained by flow cytometry of intracellular staining (**Figures S8** and **S9**). In the HTx patients, circulating CD8**^+^** T cells showed significantly higher IFNG expression compared to CD4**^+^** T cells (**Figure 6A**). GZMB and PRF were most highly expressed by NK cells.

**Figure 6.**
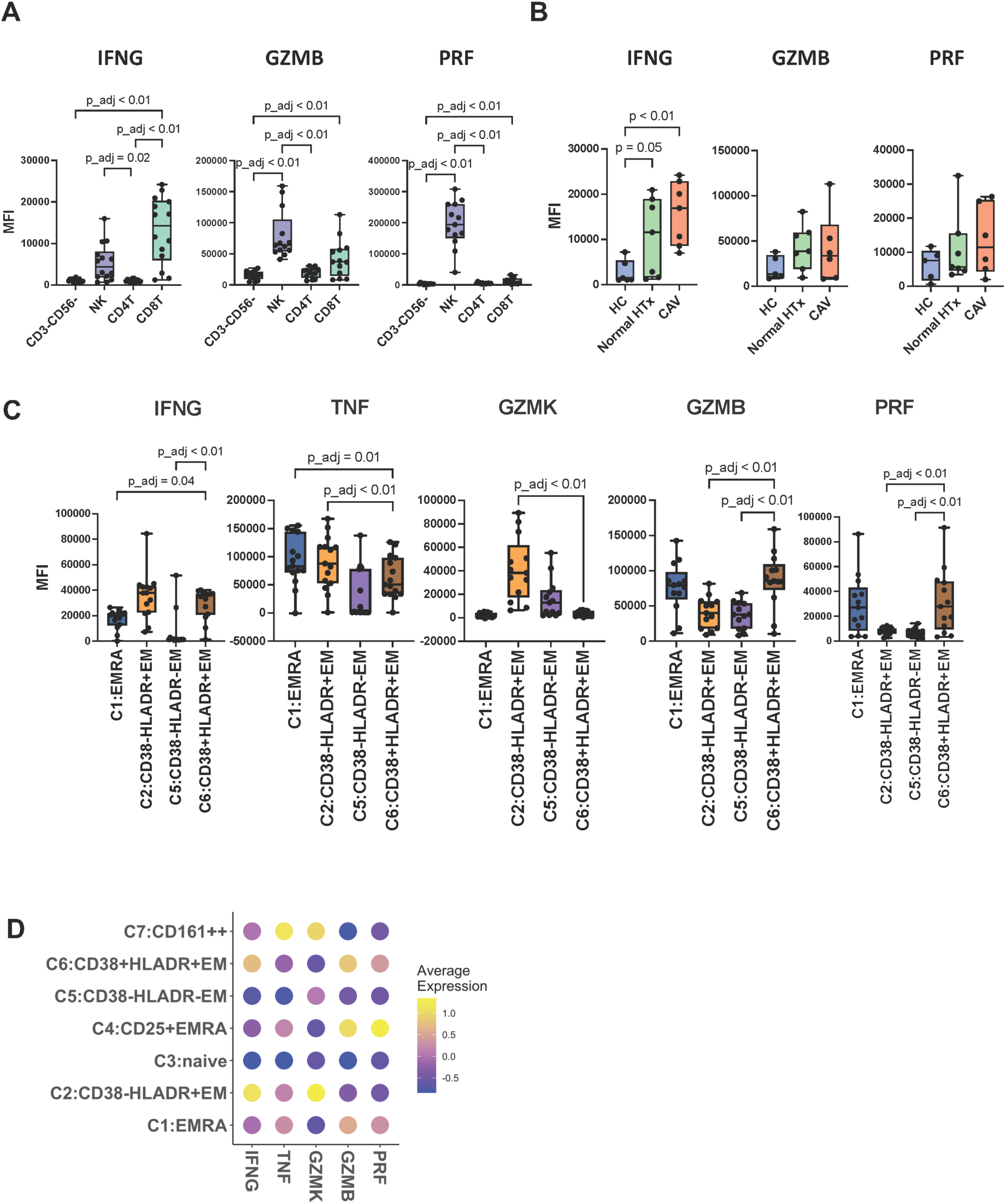
Intracellular staining demonstrates increased IFNG, GZMB, and PRF expression in circulating CD38^+^HLA-DR^+^ CD8^+^ Tem cells. **A.** Median fluorescence intensity (MFI) of intracellular staining of IFNG, GZMB, and PRF were compared among the CD3(-)CD56(-) (non-T, non-NK), NK, CD4^+^ T, and CD8^+^ T cells from the HTx patients. CD8^+^ T cells expressed higher IFNG compared to CD4^+^ T cells. **B.** Expression of IFNG, GZMB and PRF in CD8^+^ T cells are compared among the HC, normal HTx, and high grade CAV groups. High grade CAV patients showed higher IFNG expression compared to HC. **C.** Expression of intracellular markers were compared among the Temra (C1), CD38^-^HLA-DR^+^ Tem (C2), CD38^-^ HLA-DR^-^ Tem (C5), and CD38^+^HLA-DR^+^ Tem (C6) CD8^+^ T cell clusters. C6 and C2 expressed higher IFNG compared to the other clusters. C6 also expressed high levels of GZMB and PRF similarly to Temra clusters. **D.** The dot plot compares the mean values of scaled expression (z-score) of the intracellular markers among the CD8^+^ T cell clusters. For IFNG and TNF; HC (n=6), normal HTx (n=7), and high grade CAV (n=7), and for GZMK, GZMB, and PRF; HC (n=5), normal HTx (n=7), and high grade CAV (n=6). Statistical comparison was performed using a non-parametric pairwise comparison test (p-values adjusted by the Bonferroni-Holm procedure) for **A**&**C**, and Dunn test (non-adjusted) for **B**.

Circulating CD8**^+^** T cells showed increased IFNG expression in the normal HTx patient group compared to HC that was borderline significant (**Figure 6B**). High grade CAV patients expressed significantly higher IFNG MFI compared to HC. Similar trends for IFNG expression were found in circulating CD4**^+^**T cells (**Figure S9**). Expression of TNF, GZMK, GZMB, and PRF by CD8**^+^**T cells was not significantly different in high grade CAV patients compared to normal HTx or HC (**Figures 6B** and **S9**).

CD38^+^HLA-DR^+^ CD8^+^ Tem cells demonstrated significantly higher IFNG expression compared to Temra and CD38(-)HLA-DR(-) Tem (**Figure 6C, 6D**, and **S9**). CD161**^++^** CD8**^+^** T cells showed significantly higher TNF expression compared to CD38^+^HLA- DR^+^ CD8**^+^** Tem cells. CD38^+^HLA-DR^+^ CD8^+^ Tem cells demonstrated significantly higher GZMB and PRF expression compared to the other Tem clusters (C2 and C5). Thus, the increased expression of proinflammatory and cytotoxic markers by CD38^+^HLA-DR^+^ CD8^+^ Tem cells by flow cytometry corroborated the CITE-seq results (**Figures 2C** and **6D**).

### CD38^+^HLA-DR^+^ CD8**^+^** T cells were found in the intima of allograft arteries with high grade CAV

We evaluated infiltration of CD38**^+^**HLA-DR**^+^** CD8**^+^** T cells in high grade CAV (n=7) and control donor non-HTx heart tissue (n=4; **Figure 7A**). Prominent intima-media thickening with infiltration of CD4**^+^**and CD8**^+^** T cells was observed in high grade CAV but not donor heart coronary arteries. Scattered CD38**^+^**HLA-DR**^+^**CD8**^+^** T cells were also identified in the intima of the coronary arteries for four (57%) high grade CAV samples compared to none of the control samples (**Figure 7A**). In addition, five (71%) high grade CAV samples showed atherosclerotic plaques with depositions of cholesterol crystals. Ectopic lymphoid structures were also noted in the perivascular regions of all high grade CAV samples (**Figure S10**).

**Figure 7.**
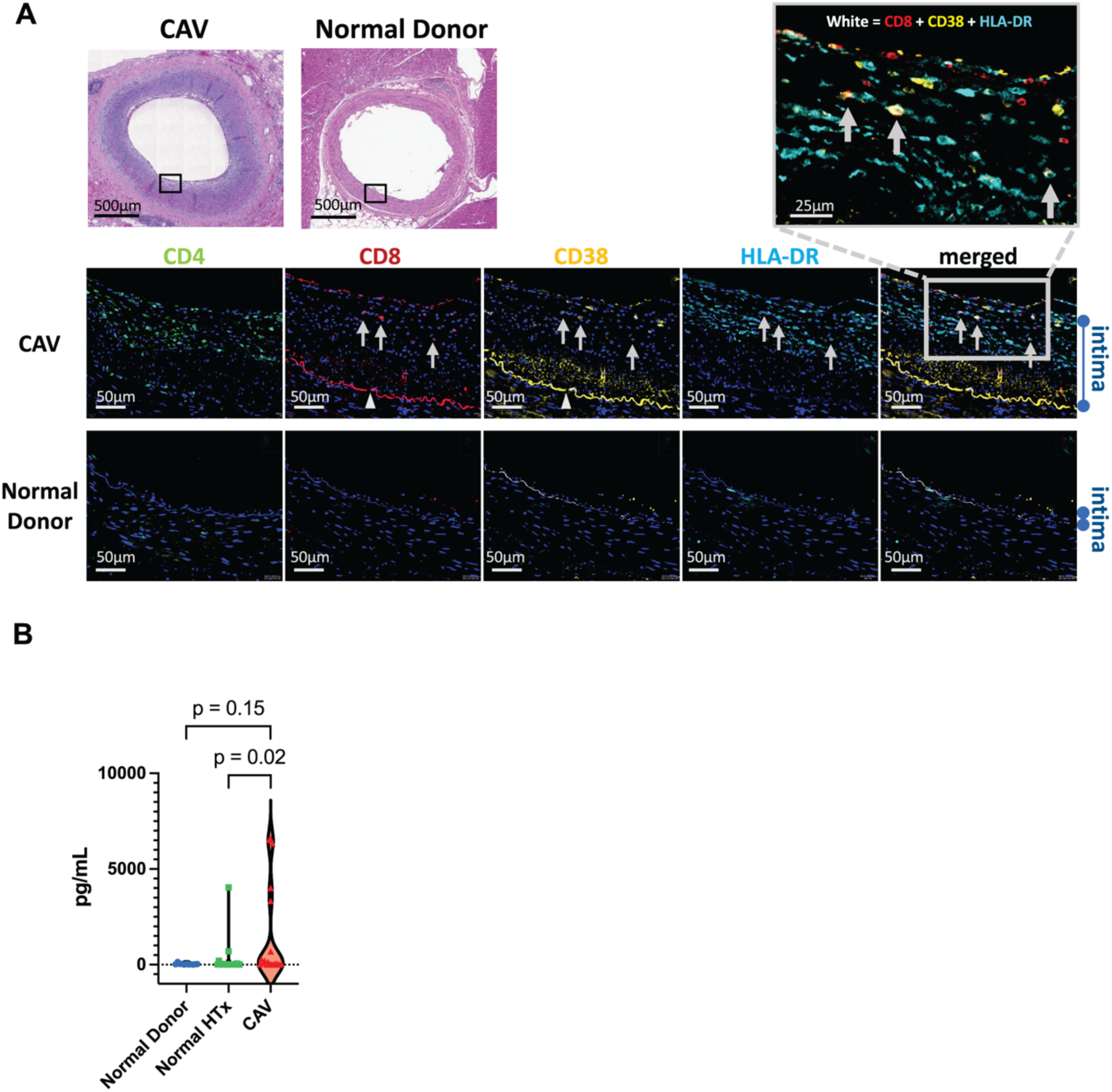
Immunopathology correlation for CD38^+^HLA-DR^+^CD8^+^ T cells and plasma LAIR2 expression. **A.** Immunofluorescence staining showed infiltration of CD38^+^HLA-DR^+^CD8^+^ T cells in the intima of allograft arteries. Top panels show low magnification images of hematoxylin and eosin staining of a high grade CAV tissue sample and a normal donor. Lower panels show immunofluorescence images of the regions of interest (black rectangles) from the hematoxylin and eosin images. Each panel shows the image CD4, CD8, CD38, HLA-DR counter-stained with Hoechst. A merged image of CD8, CD38, and HLA-DR channels is shown on the right column. Auto-fluorescence signals from the internal elastic lamina were found in the CD8 and CD38 channels (arrowheads). The high magnification merged image is shown in the right upper corner. Hoechst is excluded for visibility. CD8^+^CD38^+^HLA-DR^+^ cells are indicated by the arrows. **B.** Plasma LAIR2 levels measured by ELISA were compared among HC (n=11), normal HTx (n=20), and high grade CAV (n=20) patients (left). The high grade CAV group showed a significantly elevated plasma LAIR2 compared to normal HTx patients.

### LAIR2 expression is a potential biomarker associated with high grade CAV

Given that LAIR2 was associated with CD38**^+^**HLA-DR**^+^**CD8**^+^** T cells and significantly expressed in high grade CAV compared to normal HTx patients, we measured the plasma human LAIR2 protein level by ELISA and compared them between high grade CAV patients (n=20), normal HTx patients (n=20), and HC (n=11). Demographics of the subjects are shown in **Table S11**. High grade CAV patients showed significantly elevated plasma LAIR2 compared to normal HTx patients (70.3 pg/mL vs 16.0 pg/mL, p=0.02) (**Figure 7B**). All three patients with ISHLT grade 3 CAV who eventually required cardiac re-transplant demonstrated elevated plasma LAIR2 (>4,000 pg/mL).

## Discussion

In this study, CITE-seq demonstrated activation of peripheral CD8^+^ T cells in high grade CAV patients with an increased proportion of the CD38^+^HLA-DR^+^ CD8^+^ Tem population compared to normal HTx patients. This result was subsequently validated by flow cytometry in an independent high grade CAV patient cohort. CD38^+^HLA-DR^+^ CD8^+^ Tem cells were characterized by higher clonal expansion, activated IFNG pathway, and cytotoxicity with GZMB and PRF expression. Infiltration of the intima of CAV vessels by CD38^+^HLA-DR^+^ CD8^+^ T cells was also shown using immunofluorescence staining. Thus, our findings suggest circulating CD38^+^HLA-DR^+^ CD8^+^ Tem cells are potentially pathogenic in human CAV.

As our understanding of the CD8^+^ T cell repertoire in humans has grown increasingly complex,^19^ we utilized CITE-seq to resolve the CD8^+^ T cell diversity at a single cell resolution in high grade CAV patients. In prior animal studies, CD8^+^ T cells have been shown to cause CAV lesions, even in the presence of calcineurin inhibition.^4,5^ However, conventional flow cytometric techniques limit the initial study of peripheral CD8^+^ T cell clusters potentially contributing to human CAV. Thus, using our novel approach of first performing comprehensive cellular and transcriptomic profiling of PBMCs and subsequently validating our findings with flow cytometry, we were able to identify CD38^+^HLA-DR^+^ CD8^+^ Tem cells as a potentially pathogenic circulating CD8^+^ T cell cluster associated with high grade CAV in patients.

This is the first study to demonstrate circulating CD38^+^HLA-DR^+^ CD8^+^ Tem cells may be an important contributor to the IFNG axis in human CAV. CD38^+^HLA-DR^+^ CD8^+^ T cells have also been reported in other proinflammatory disease states. Fernandez et al previously described proinflammatory CD38^+^HLA-DR^+^ CD8^+^ T cells in native carotid atherosclerotic plaques in non-HTx patients.^20^ Co-expression of CD38 and HLA-DR in peripheral CD8^+^ T cells have also been shown in HIV and hepatitis C non-HTx patients to be associated with increased IFNG production.^21,22^ While prior studies have suggested that the IFNG axis is central to the development of human CAV,^8,23^ the contribution of peripheral T cells to the IFNG axis has not been fully studied in human CAV. Thus, our study findings are critical to establish the link between proinflammatory circulating CD38^+^HLA-DR^+^ CD8^+^ Tem cells and the IFNG axis in human CAV.

In addition to the proinflammatory phenotype, we also observed expression of cytotoxic cytokines by circulating CD38^+^HLA-DR^+^ CD8^+^ Tem cells. Prior studies suggest that both cytotoxic and proinflammatory pathways mediated by CD8^+^ T cells are necessary for CAV development.^7,24^ Choy et al. have previously demonstrated that GZMB works together with PRF to induce endothelial cell apoptosis that subsequently leads to CAV development.^24^ In our study, we observed infiltration of CD38^+^HLA-DR^+^ CD8^+^ T cells in the intima of human allograft arteries with high grade CAV. Thus, we hypothesize that circulating CD38^+^HLA-DR^+^ CD8^+^ Tem cells may infiltrate intima of allograft arteries to cause intimal injury through proinflammatory and cytotoxic pathways, ultimately leading to CAV.

Our findings also demonstrated significant clonal expansion of CD8^+^ T cells in high grade CAV compared to normal HTx patients. In particular, oligoclonality of the CD38^+^HLA-DR^+^ CD8^+^ Tem cells suggest that this cluster is expanded through chronic antigen stimulation and is not a result of a bystander effect. In addition, we found that the expanded clonotypes of CD38^+^HLA-DR^+^ CD8^+^ Tem cells were shared by both Temra and CD8^+^ Tem cells. We also demonstrated that the diffusion pseudo-time of CD38^+^HLA-DR^+^ CD8^+^ Tem cells were intermediate to Tem and Temra cell clusters. Thus, these data support our hypothesis that CD8^+^ Tem cells expand and differentiate into CD38^+^HLA-DR^+^ CD8^+^ Tem and eventually CD8^+^ Temra cells.

Another novel finding observed in our study was the differences of CD38 and HLA-DR expression in relation to CD8^+^ T cell differentiation. While CD38 and HLA-DR expression are both considered activation markers, CD38 and HLA-DR expression patterns in association with CD8^+^ T cell differentiation have not been previously described in patients. In our study, HLA-DR expression initially increased with CD8^+^ T cell differentiation status, with peak expression in CD38^+^HLA-DR^+^ Tem cells, before a subsequent decline seen with Temra cells. In contrast, CD38 expression first decreases with CD8^+^ T cell differentiation status before increasing again in the more differentiated CD38^+^HLA-DR^+^ Tem and Temra clusters. Indeed, human CD38 is a complex, multifunctional glycoprotein that is also a marker for T cell overactivation or exhaustion.^25^ This may also explain our observation that the most differentiated CD8^+^ Temra cells were CD38 positive but HLA-DR negative.

While we have demonstrated that CD38^+^HLA-DR^+^ CD8^+^ Tem cells are associated with high grade CAV in patients, we hypothesize that CAV is likely a multifactorial disease where antibody mediated rejection (AMR) and T cell-mediated immunity may contribute in varying degrees for different CAV patients. Clinical studies have shown that persistent donor specific anti-HLA antibody (DSA) production and AMR development are associated with greater risk of CAV.^26,27^ Allorecognition by CD4^+^ T cells in chronic rejection has also been shown to contribute to development of DSAs and AMR.^28^ In the current study, there was no significant difference in AMR or DSA prevalence in the high grade CAV compared to the normal HTx patient group for both CITE-seq and flow cytometry patient cohorts. In addition, we did not detect any significant differences in circulating CD4^+^ T cell clusters in the high grade CAV compared to normal HTx patients and circulating CD8**^+^** T cells showed significantly higher IFNG expression compared to CD4**^+^** T cells. However, we did observe ectopic lymphoid structures perivascular regions of high grade CAV samples, introducing the possibility of local alloantibody production contributing to CAV development.^29^ Additional research, beyond the scope of this work, is needed to explore the relationship between CD38^+^HLA-DR^+^ CD8^+^ Tem cells and high grade CAV patients in the context of AMR and/or ectopic lymphoid structures.

As the mechanisms of CD38^+^HLA-DR^+^ CD8^+^ Tem cells contributing to CAV are further defined, we hypothesize that identification of circulating CD38^+^HLA-DR^+^ CD8^+^ Tem cells themselves could be a potential early biomarker to predict future high grade CAV in patients. With a panel of limited surface marker antibodies, we show it is feasible to identify CD38^+^HLA-DR^+^ CD8^+^ Tem cells from PBMCs using flow cytometry. This may eventually be translated to a clinical laboratory setting using whole blood samples. However, whether CD38^+^HLA-DR^+^ CD8^+^ Tem cells predict development of high grade CAV and also perhaps earlier CAV grades will require future longitudinal studies to demonstrate clinical validity.

Lastly, we found LAIR2 as a promising protein biomarker for identifying high grade CAV patients. LAIR1 has been shown to negatively regulate immune cell function, such as inhibition of Th1 immunity.^30^ LAIR2 is less well known and is thought to activate immunity by inhibiting the LAIR1 pathway.^31^ In our CITE-seq cohort, LAIR2 expression in CD8^+^ T cells was significantly increased in high grade CAV compared to normal HTx patients. However, we were able to go further and demonstrate significantly increased LAIR2 plasma levels in high grade CAV compared to normal HTx patients in an independent cohort. Thus, the potential for plasma LAIR2 as a noninvasive test to predict high grade CAV associated with poor prognosis is appealing and warrants further exploration, particularly given the current lack of a simple blood test to identify this critical group of patients.^32^

### Strengths and Limitations

First, while our study is inherently limited by the small number of patients examined, the strength of our study is that the key findings in the CITE-seq cohort were validated in a larger independent patient cohort using conventional flow cytometry. Additionally, clusters determined by CITE-seq and flow cytometry demonstrated comparable expression patterns of inflammatory cytokines and cytotoxic enzymes. Thus, our CITE-seq and corroborating flow cytometry results provide confidence in the clinical validity of our study findings. Second, as CAV can be associated with different clinical risk factors,^33,34^ we sought to evaluate the CD8^+^ T cell response in high grade CAV patients without a hepatitis C nucleic acid test positive donor heart or previous HTx to reduce the potential confounders affecting peripheral immune cell response interpretation. Consequently, our findings may not necessarily be representative of all HTx patients that develop high grade CAV. Third, since we did not evaluate patients with ISHLT grade 1 CAV, it is possible that our findings may not represent the immune response during the earlier stages of CAV. However, we believe our study findings provide the necessary rationale for future longitudinal studies that can employ flow cytometry to identify the potentially pathogenic CD38^+^HLA-DR^+^ CD8^+^ Tem cells to investigate early CAV. Fourth, access to tissue that allows for detailed characterization of human cardiac allograft arteries from normal HTx patients as a comparator to explanted CAV allografts is a recognized limitation in this field of study.^35^ Thus, the focus of our study was on the evaluation of PBMCs as these samples are accessible for both CAV and normal HTx patient cohorts and the research findings can be more readily translated into the clinical setting.

### Conclusion

We conclude that PBMCs from high grade CAV patients demonstrate significantly expanded CD38**^+^**HLA-DR**^+^** effector memory CD8**^+^** T cells with increased IFNG signaling and cytotoxicity compared to normal HTx patients. Detection of circulating CD38**^+^**HLA-DR**^+^**effector memory CD8**^+^** T cells and LAIR2 are promising candidate biomarkers that may predict high grade CAV.

## Acknowledgement

The authors acknowledge Helen Smith, PhD, at BD Bioscience for providing instructions and technical advice for using the BD Rhapsody single-cell analysis system and related sequencing, Suzie Alarcon, BS, and the LJI sequencing core for assistance with sequencing our samples, the LJI histology core for their assistance with immuno-staining of our tissue samples, Vincenzo Cusi, BS, for his assistance with coordinating sample collections from patients, and Parker Dow, BS, for his contribution to our T cell receptor analyses.

## Supplemental Material

Table S1-11

Figure S1-10

Supplemental Excel File 1-5

References #36-#40

## Non-standard Abbreviations and Acronyms

ACR: acute cellular rejection
ADT: antibody-derived tag
AMR: antibody mediated rejection
CAV: cardiac allograft vasculopathy
CITE-seq: cellular indexing of transcriptomes and epitopes by sequencing
DC: dendritic cells
DSA: donor specific anti-HLA antibodies
GSEA: gene set enrichment analysis
GZMB: granzyme B
GZMK: granzyme K
HLA: human leukocyte antigen
HTx: heart transplant
ICS: intracellular staining
IFNA: interferon-α
IFNG: interferon-γ
ISHLT: International Society of Heart and Lung Transplant
LAIR: leukocyte-associated immunoglobulin-like receptor
LVEF: left ventricular ejection fraction
MFI: median fluorescence intensity
NK: natural killer
PBMC: peripheral blood mononuclear cell
PRF: perforin
TCR: T cell receptor
Tem: effector memory
Temra: effector memory re-expressing CD45RA
TNF: tumor necrosis factor
UCSD: University of California, San Diego
UMAP: uniform manifold approximation and projection
UMI: unique molecular identifiers
WNN: weighted-nearest neighbor

